# Modelling the impact of interventions on the progress of the COVID-19 outbreak including age segregation

**DOI:** 10.1101/2020.04.04.20053017

**Authors:** Jorge Rodríguez, Mauricio Patón, Joao M Uratani, Juan M Acuña

## Abstract

Infectious diseases can be devastating, especially when new and highly contagious, producing epidemic outbreaks that can become pandemics. Such is the case of COVID-19, the worst pandemic the world has seen in more than 100 years. Predicting the course and outcomes of such a pandemic in relation to possible interventions is crucial for societal and healthcare planning and forecasting of resource needs. In this work a deterministic model was developed, using elements from the SIR-type models, that describes individuals in a population in compartments by infection stage and age group. The model assumes a close well-mixed community with no migrations. Infection rates and clinical and epidemiological information govern the transitions between stages of the disease. The present model provides a platform to build upon and its current low complexity retains accessibility to both experts and non-experts as well as policy makers to comprehend the variables and phenomena at play.

The impact of several possible interventions that have been or may be applied to slow the spread of the COVID-19 outbreak is evaluated. Key findings in our model simulation results indicate that (i) universal social isolation measures may be effective in reducing total fatalities only if they are strict and the average number of daily social interactions is reduced to very low numbers; (ii) selective isolation of only the age groups most vulnerable to the disease (i.e. older than 60) appears almost as effective in reducing total fatalities but at a much lower economic damage; (iii) the use of protective equipment (PPE) appears capable of very significantly reducing total fatalities if implemented extensively and to a high degree; (iv) extensive random testing of the population leading to infection recognition and subsequent immediate (self) isolation of the infected individuals, appears to be an ineffective intervention due to the required (unreachable with existing test sensitivities) high percentage of infection detections and the incapability to be sustained over time; (v) an increase in the number of critical care beds to directly save significant numbers of lives with a direct reduction in total final fatalities per each extra available critical care bed unit.

## Introduction

Mathematical modelling of the spread of diseases has a long history. Epidemiology models can be either deterministic or stochastic. Deterministic compartmental models (DCM) known as SIR (Susceptible-Infected-Recovered) models were published in the early 20^th^ century (Kermack & McKendrick, 1927). The acronym of the model is based on which compartments of people are included within the model. Developments of the SIR models have resulted in different variations such as SEIR/MSEIR/SEIQJR and other compartmental models (Hethcote, 2000). Deterministic epidemic models allow for the understanding of the population-based dynamics as well as for the identification of parameters of interest (e.g. transmission rates). Numerous examples of deterministic SIR (or variations of SIR) models have been published in the literature (May & Anderson, 1979; Ruan & Wang, 2003; Li *et al*., 2004).

Alternative modelling approaches have focused on the randomness nature of the spread of a disease, particularly at the start of an outbreak, when a low number of people are infected. Therefore, stochastic modelling can be an adequate approach to model such spread events (Britton, 2010). Most of the stochastic models for spread diseases have evolved into network models (McCallum *et al*., 2001; Keeling & Eames, 2005; Small and Tse, 2005; Ferguson et. al. 2006; Grassly & Fraser, 2008; Keeling & Rohani, 2008; Balcan *et al*., 2010; Gray *et al*., 2011; Brauer *et al*., 2012; Miller *et al*., 2012; Siettos & Russo, 2013; Pastor-Satorras *et al*., 2015). In any case, the outputs of the epidemic models are frequently used to inform studies on health projections and play an important role in shaping policies related to public health (Murray and Lopez, 1997a, 1997b, 1997c, and 1997d; Ferguson *et al*., 2006).

Data timelines and availability has greatly increased in recent years, which led to direct improvements in epidemic models (Colizza *et al*., 2006; Riley, 2007; Siettos & Russo, 2013). These models can help in providing a more comprehensive understanding of recent outbreaks of diseases such as Ebola (Gomes *et al*., 2014; WHO Ebola Response Team, 2014) and Zika (Zhang *et al*., 2017). However, all modelling efforts are highly dependent on several elements: a deep understanding of the course of the disease; a comprehensive algorithm of clinical and public health options available and stages of events; probability of such options given certain conditions of the system; identification of parameters that reflect such events and their probabilities (such as mortality by age, infectiousness by contacts, etc.); best assumptions for parameters with insufficient data; and valid data for those parameters that allow the calibration and posterior validation of the forecasts (Tizzoni *et al*., 2012). In order to obtain a proper and timely identification of parameters for the models, access to up-to-date data is required, in this case of the COVID-19 spread (Dong *et al*., 2020).

In viral pandemics in particular, one of those parameters, the direct estimation of infected sub-population fractions, is not feasible using available epidemiological data (unless universal, highly sensitive testing is used, which is rarely possible to implement in these situations), particularly if very mild cases, asymptomatic infections or pre-symptomatic transmission are observed or expected. This was the case of the previous Influenza A (H1N1-2009) pandemic (Nishiura *et al*., 2011) and it is the observation for the COVID-19 pandemic (Russell *et al*., 2020). Thus, in many cases, modelling uses a combination of the best available data from historical events and datasets, parameter estimation and assumptions. Then, data about these parameters are computed with statistical tools for the identification of parameters that are used in epidemic models (Cooper *et al*., 2006; Biggerstaff *et al*., 2014).

The most challenging cases for the understanding of the potential spread of a disease is when a novel disease outbreak emerges in global populations (Anderson & May, 1992). The characteristics of the novel disease (e.g.: R_0_, fatality rate and the clinical course of the disease) are unknown as the data availability is limited (e.g. novelty of pathogen; delay of communication of case datasets from public health workers and facilities to researchers) or biased (*e*.*g*., limited availability of testing capacity; undefined or partially defined diagnostics for disease). With novel disease-specific epidemic models, the development of models as low complexity as possible but with as many meaningful parameters as needed, is paramount. These parameters can be accurately defined with data as the infection progresses and data become more available, and the model may be posited as a potential tool to inform public health policy and impact mitigation strategies (Berezovskaya *et al*., 2005; Hall *et al*., 2007; Bettencourt *et al*., 2008; Nishiura, 2011; Wang & Zhao, 2012; Lee *et al*., 2013; Nsoesie *et al*., 2014, Chowell *et al*., 2016, Rivers *et al*., 2019, Chowell *et al*., 2020).

The COVID-19 outbreak and posterior pandemic has brought unprecedented attention into these kinds of modelling and its limitations, with multiple epidemic models and disease spread forecasts being published as more data becomes available. These models have evaluated the ongoing course of the disease spread evolution, from the earlier dynamics of transmission from initial cases (Kucharski *et al*., 2020), to the potential of non-pharmaceutical interventions to limit the disease spread, such as: international travel restrictions (Chinazzi *et al*., 2020), contact tracing and isolation of infected individuals at onset (Hellewell *et al*., 2020), different scales of social distancing and isolation (Flaxman *et al*., 2020, Prem *et al*., 2020). Other statistical models tried to estimate fundamental characteristics (*i*.*e*. potential model parameters) for the disease, such as the incubation period (Lauer *et al*., 2020) as well as to assess short-term forecasts (Roosa *et al*., 2020). Given the inherent uncertainty associated with most of the parameters used, stochastic approaches are employed in the above models.

Effective communication between health care and public health systems and science hubs is considered one of the bigger challenges in both health sciences and public health (Zarcadoolas, 2010; Squiers *et al*., 2012). In health care It is not only necessary to take effective measures but also to do it timely. This requires strategies for data sharing, generation of information and knowledge and timely dissemination of such knowledge for effective implementations. User-accessible modelling tools can contribute to the understanding by broader audiences (researchers, public health authorities, and the general public) of what to expect on the propagation of infectious diseases and how specific interventions may help. This increased awareness of the disease behaviour and potential course in time by public and policy makers can directly and positively impact the outcome of epidemic outbreaks (Funk *et al*., 2009).

The present work presents a fully deterministic SIR-type model aimed at the evaluation of intervention scenarios for the COVID-19 outbreak. The model introduces the novel approach of compartmentalisation by age groups and known disease stages, crucial and specific level of complexity for COVID-19 prediction. This aims at retaining mechanistic meaning of all variables and parameters while capturing the relevant phenomena at play. The complexity level was limited specifically to maintain accessibility to non-experts and policy makers to comprehend the model results such that expert advice and decision making can be brought closer together to help guide interventions for immediate and longer-term needs.

## Model description

The model presented is based on balances of individuals, segregated by age group, transitioning between infection stages. All individuals are placed in a common single domain or closed community (e.g. a well-mixed city or town), no geographical clustering nor separation of any type is considered, nor is any form of migration in or out of the community. Large cities with ample use of public transportation are thought to be settings best described by the model.

The model also provides a direct estimation of the dynamic reproduction number or reproductive rate (R_t_) (Delamater *et al*. 2019; De Serres *et al*. 2000; Lihong *et al*. 2020) under different circumstances of individual characteristics (such as use of personal protection or awareness) as well as under population-based interventions (such as imposed social isolation). R_t_ is a dynamic number often quoted erroneously as a constant for a specific microorganism or disease. The ability to estimate the R_t_ for different times of the outbreak (given the interventions), outbreak settings and interventions is considered to be a valuable characteristic of the model. R_t_ is predicted to change over time with interventions not related to increased immunity (isolation or use of personal protection equipment (PPE), as opposed to vaccination).

### Model constituents

The model solves dynamic variables or states. Every individual belongs, in addition to their age group (which she/he never leaves), to only one of the possible states that correspond to stages of the infection in terms of infectiousness and severity of symptoms, namely: *healthy susceptible* (H); *pre-symptomatic non-infectious* (NI); *pre-symptomatic infectious* (PS); *symptomatic* (S); *in need of hospitalisation* (SH); *in need of critical care* (SC); *deceased* (D) and *recovered immune (*R).

Definitions of the model state variables are shown in Table 1. Each variable is a state vector with the number of individuals in that stage per age group, which is defined per decade from 0-9 to 80+ year olds (9 age groups). Therefore, each state is a vector of dimensions 1×9, and the total number of states is a matrix of dimensions 8×9. Note that vector variables and parameters are represented in bold font and scalar ones in regular font.

**Table 1.** Model states in vectors (1×9) of number of individuals in each infection stage.

| Definition “Number of individuals...” | Variable | Totals of all ages |
| --- | --- | --- |
| Healthy susceptible to infection | **$N_h$** | $N_{hT}$ |
| Non-infectious pre-symptomatic | **$N_{ni}$** | $N_{niT}$ |
| Infectious pre-symptomatic | **$N_{ps}$** | $N_{psT}$ |
| Infectious symptomatic | **$N_s$** | $N_{sT}$ |
| Requiring hospitalisation | **$N_{sh}$** | $N_{shT}$ |
| Requiring critical care | **$N_{sc}$** | $N_{scT}$ |
| Deceased | **$N_d$** | $N_{dT}$ |
| Recovered & immune | **$N_r$** | $N_{rT}$ |

Figure 1 shows the population progress through stages as described in the model. An additional schematic representation of the model approach with the population groups considered for the infection stages, rates of infection and transition between groups and showing possible interactions between population groups is shown in Figure S5.

**Figure 1.**
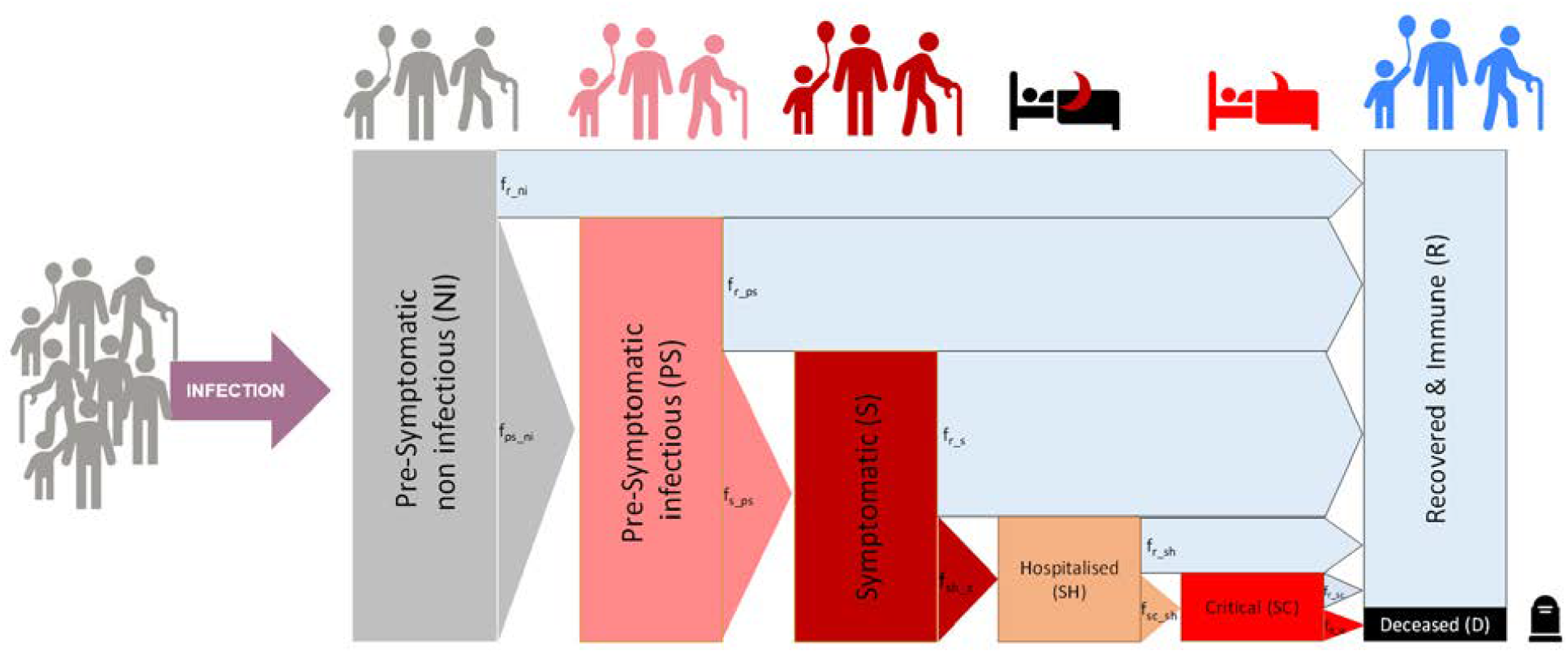
Schematic representation of the population compartments by infection stages and age. Only interactions between infectious individuals (both pre-symptomatic and symptomatic) and healthy susceptible ones can increase the rate of infections.

### Rates of transition between infection stages

The transitions between stages are governed by the rates of infection and transition shown in Table 2.

**Table 2.**
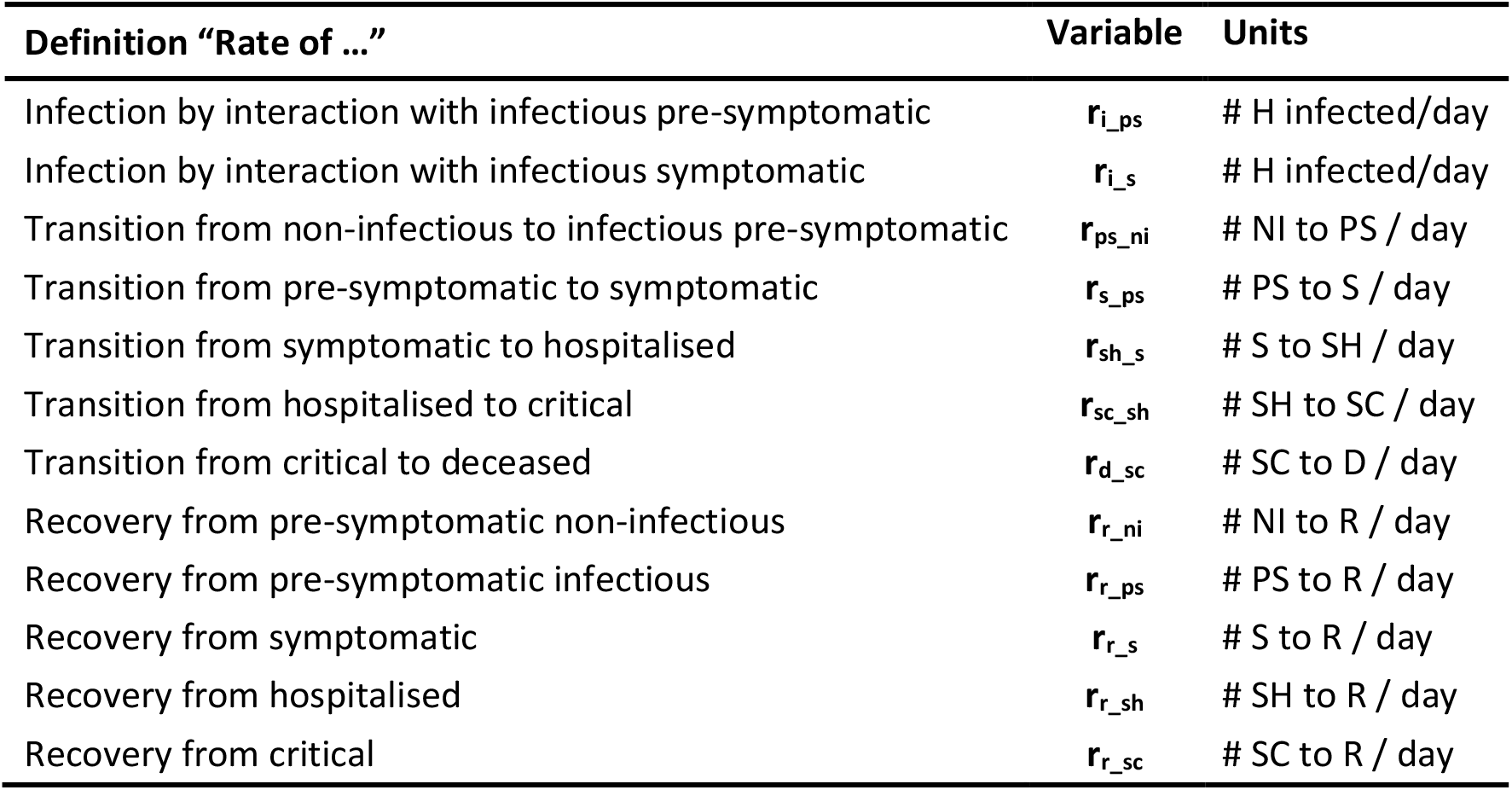
Rates of infection and transition between states, vectors (1×9).

The average rates of transition between states are defined such that the latest epidemiological and clinical data can be used to determine and continuously update the parameters as more knowledge of the disease emerges. These parameters include the proportion of individuals that transition to a more severe stage or recover (see Table 3) and the average times reported at each stage before transition or recovery (see Table 4).

**Table 3.** Epidemiological parameters (all in vectors per age group).

|  |  |  |  |
| --- | --- | --- | --- |
| Fraction of PS that will become S | | $f_{s\_ps}$ | #S/#PS |
| Fraction of S that will become SH | | $f_{sh\_s}$ | #SH/#S |
| Fraction of SH that will become SC | | $f_{sc\_sh}$ | #SC/#SH |
| Fraction of cared SC that will die into D | | $f_{d\_sc}$ | #D/#SC<sub>IC</sub> |
| Fraction of NI that will recover into R<sup>1</sup> | $(1 - f_{ps\_ni})$ | $f_{r\_ni}$ | #R/#NI |
| Fraction of PS that will recover into R<sup>1</sup> | $(1 - f_{s\_ps})$ | $f_{r\_ps}$ | #R/#PS |
| Fraction of S that will recover into R<sup>1</sup> | $(1 - f_{sh\_s})$ | $f_{r\_s}$ | #R/#S |
| Fraction of SH that will recover into R<sup>1</sup> | $(1 - f_{sc\_sh})$ | $f_{r\_sh}$ | #R/#SH |
| Fraction of cared SC that will recover into R<sup>1</sup> | $(1 - f_{d\_sc})$ | $f_{r\_sc}$ | #R/#SC<sub>IC</sub> |

<sup>1</sup>Calculated by difference with the complementary, not an input parameter

**Table 4.** Clinical average times in each infection stage (all in vectors per age group).

| Definition | Parameter | Units |
| --- | --- | --- |
| Time to become infectious | $t_{ps\_ni}$ | days |
| Time to develop symptoms from infectious | $t_{s\_ps}$ | days |
| Time to require hospitalisation from symptoms onset | $t_{sh\_s}$ | days |
| Time to require critical care from hospitalisation | $t_{sc\_sh}$ | days |
| Time to death from critical | $t_{d\_sc}$ | days |
| Time to death from critical with no care available | $t_{d\_nc}$ | days |
| Time to recover from presymptomatic non-infectious | $t_{r\_ni}$ | days |
| Time to recover from presymptomatic infectious | $t_{r\_ps}$ | days |
| Time to recover from (non-severe) symptoms | $t_{r\_s}$ | days |
| Time to recover from hospitalisation | $t_{r\_sh}$ | days |
| Time to recover from critical | $t_{r\_sc}$ | days |

The rates of transition between stages (in number of individuals per day) are described in Eqs. 1.a-f. All rates are vectors per age group of dimensions (1×9). Note that point operators between vectors indicate an operation element-by-element.

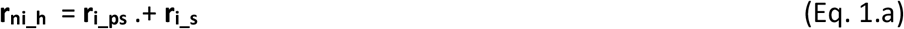

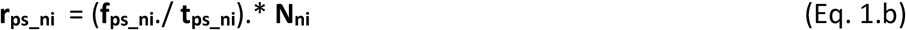

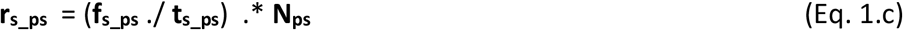

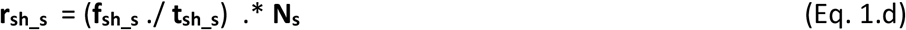

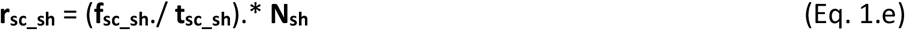

The rates of individuals recovering from the different infected stages (in number of individuals per day) are described in Eqs 2.a-e. (all rates in vectors per age group).

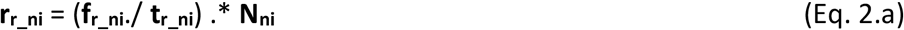

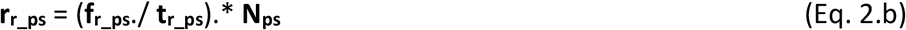

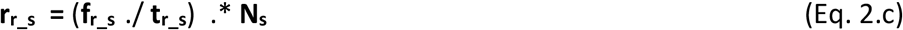

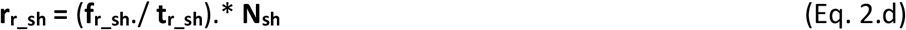

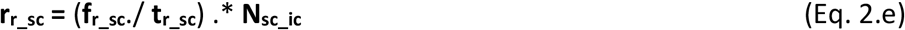

The rate of transition from critical to deceased is the sum of that of those critical receiving intensive care (**r**_**d_scic**_) plus that of those critical without available care (**r**_**d_scnc**_) as per Eqs. 3.a-c. All critical individuals not receiving intensive care (**N**_**sc_ncc**_) are assumed to become fatalities after a time (**t**_**d_nc**_). The allocation of critical care is described below.

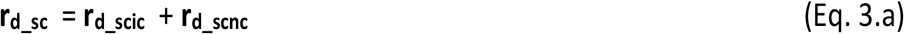

where

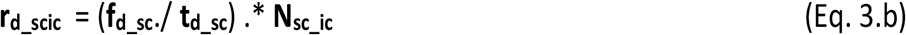

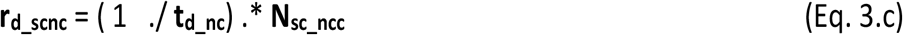

Figure 2 provides a representation of the transition between stages, the proportions of individuals recovering or worsening are as per Figure 1.

**Figure 2.**
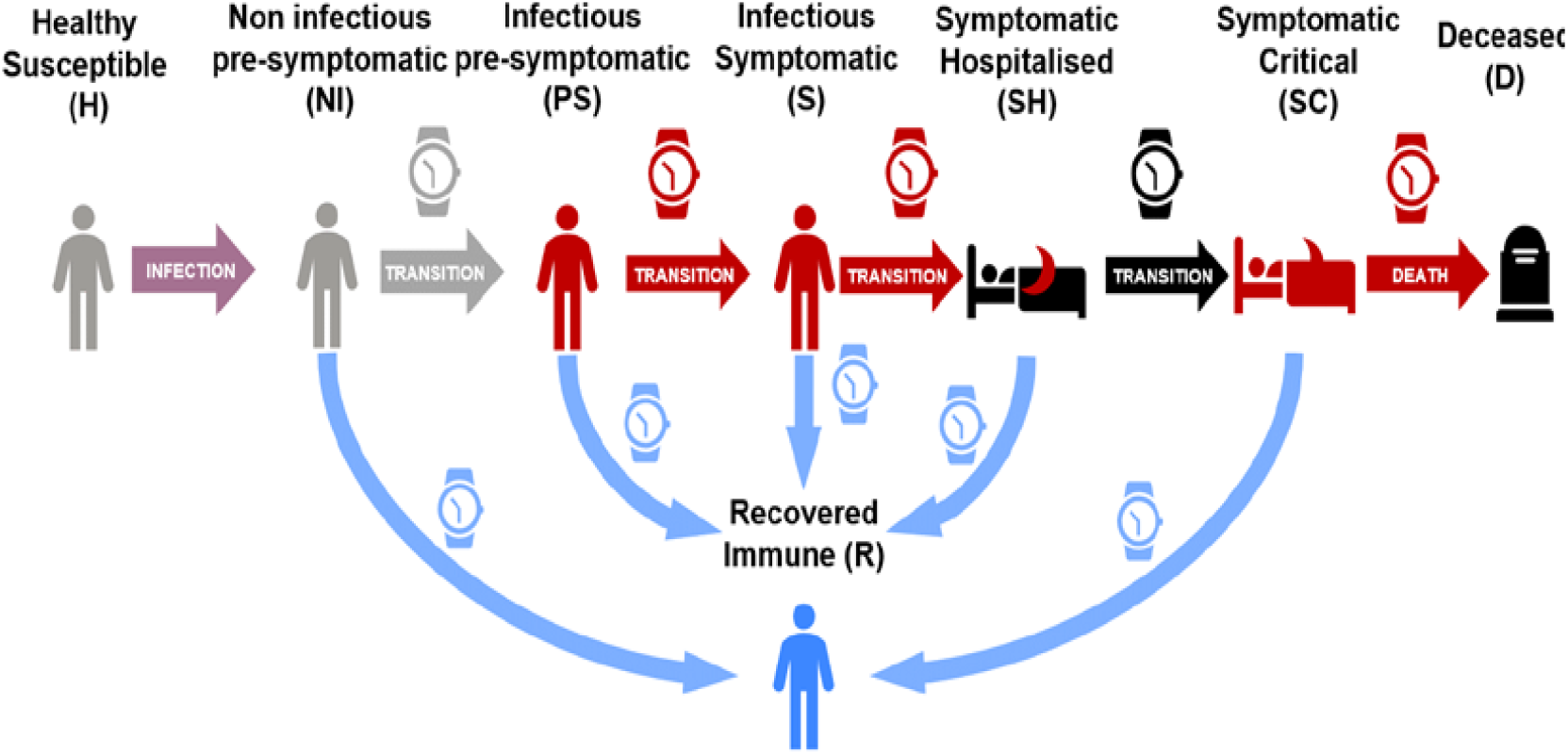
Schematic representation of the transitions rates between stages. Individuals spend in each stage an amount of time depending on their transition path towards recovery or increased severity.

### Rates of infection

The infection of healthy susceptible individuals (H) is modelled as occurring only via their interaction with infectious either pre-symptomatic (PS) or symptomatic (S) individuals. Hospitalised (SH) and critical (SC) individuals are assumed not available for interactions neither are those deceased (D).

Two rates of infection of healthy susceptible individuals (in number of infections per day) are defined, one from each one of the two possible infecting groups (PS and S). The rates of infection (in vectors per age group) result from the product of (i) the fraction of interactions occurring with PS (or S) individuals among the total interactions (*f*_ips_ or *f*_is_) times (ii) the likelihood of contagion in an interaction with PS (or S) (**p**_**i_ps**_ or **p**_**i_s**_) (per age group), (iii) the average number of daily interpersonal contacts that H individuals have (**ni**_**h**_) and (iv) the number of H individuals themselves (per age group) (see Eqs 4.a-b and Figure 3).

**Figure 3.**
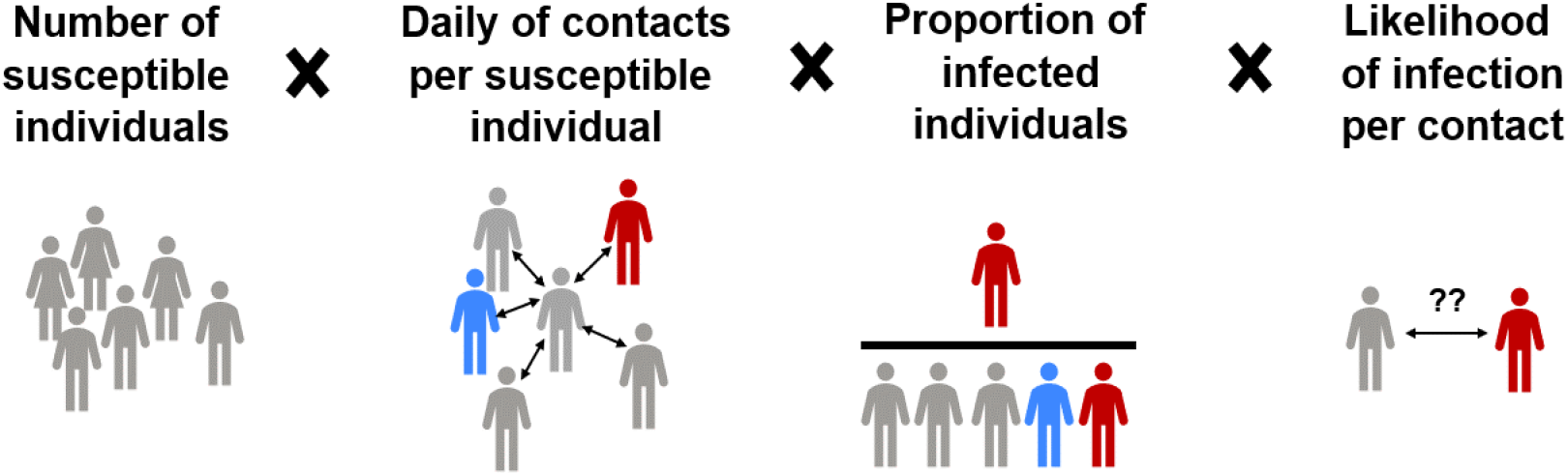
Schematic representation of the terms of the infection rate.

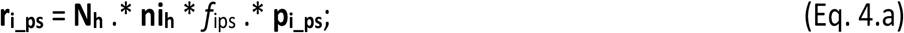

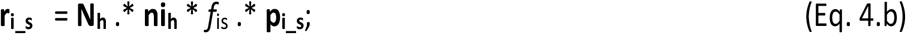

### Modelling interventions

Four main possible interventions are evaluated that have been or may be applied to slow the spread of the COVID-19 outbreak namely (i) the *degree of social isolation* of the individuals in the population, in terms of the average number of interpersonal contacts that individuals have per day with other interacting individuals; (ii) the *level of personal protection and awareness* that individuals apply to protect themselves and others against contagion during interactions; (iii) the *percentage of infected-aware individuals* due to testing that will subsequently quarantine and (iv) the intensive care capacity available. These interventions can be stratified by age groups. Table 5 describes the key parameters that define the interventions.

**Table 5.**
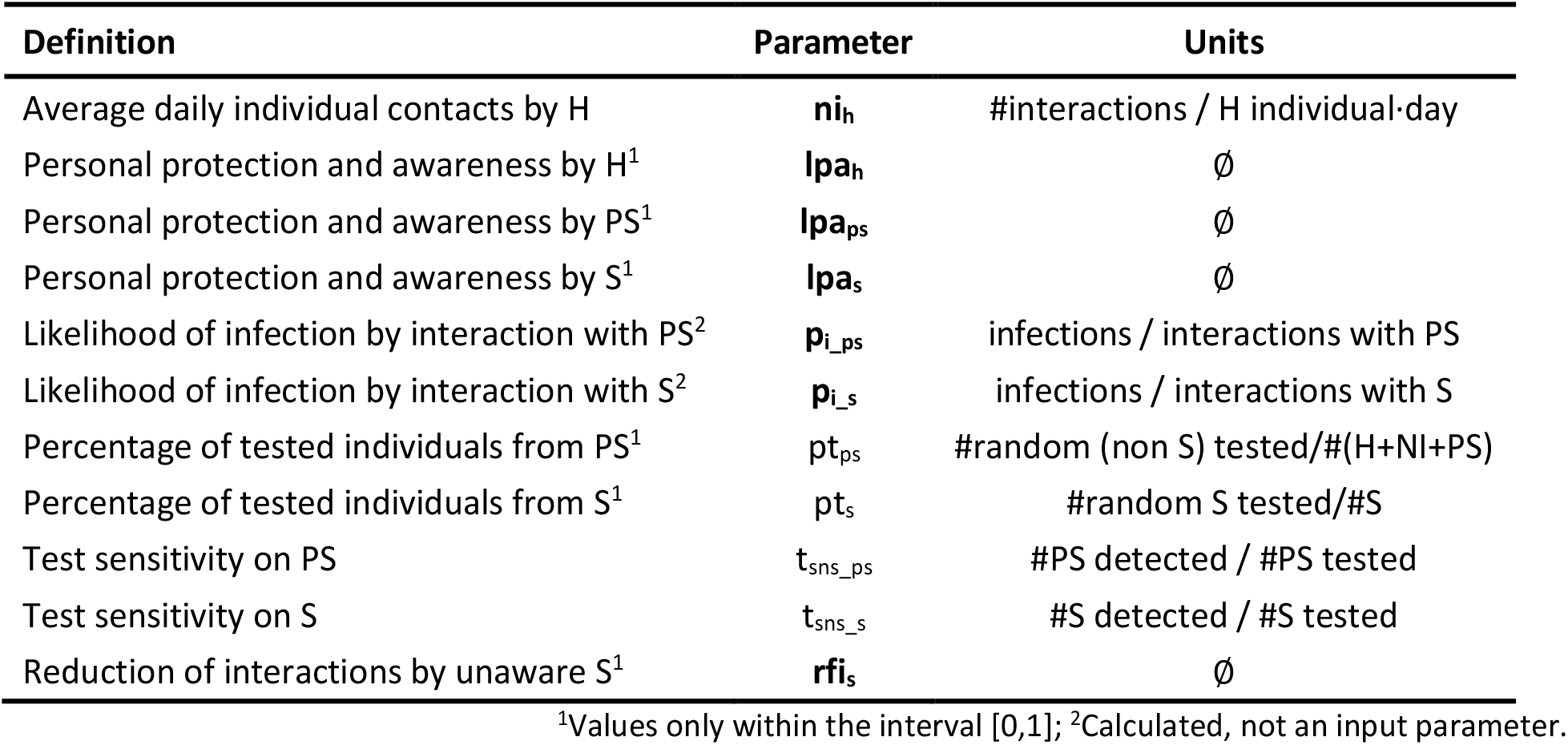
Intervention parameters

#### Social isolation

The *degree of social isolation* is described through its impact on the parameter (**ni**_**h**_) (vector per age group) corresponding to the representative average number of interpersonal contacts that healthy susceptible individuals have with others per day. Different **ni**_**h**_ values can be applied to different age groups to describe age selective isolation strategies such as e.g. isolation of the elderly and/or the young alone.

#### Use of PPE and safety distance

The *level of PPE use and awareness* is described as impacting the likelihood of infection (see Eqs. 4) through the parameters (**lpa**_**h**_) for healthy and (**lpa**_**ps**_ and **lpa**_**s**_) for infectious PS and S individuals (both in vectors per age group). Values of the **lpa** parameters can vary between 0 and 1, with 1 corresponding to the use of complete protective measures and zero to the most reckless opposite situation. Different values can be assigned e.g. for children and adults as well as for those suffering from symptoms (S).

The likelihoods of infection per interaction are calculated as per Eq. 5.a-b.

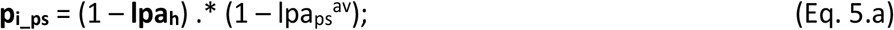

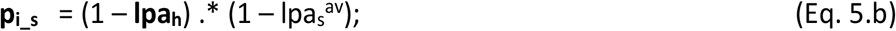

where **lpa**_**h**_ reflects a level of protection and awareness related to interventions and defined below; Ipa_as_^av^ and Ipa_s_^av^ are scalars corresponding to the weighted averages over all age groups of the pool of PS and S with which H individuals can interact (Eqs 5.c-d).

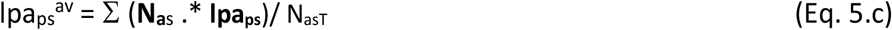

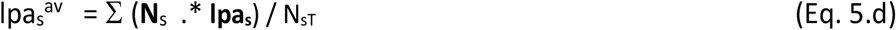

where N_psT_ and N_sT_ are the total numbers of PS and S individuals of all ages respectively. The ∑ symbol indicates summation across all age groups.

#### Awareness of infection by testing

The *awareness of infection* after positive testing is described through a reduction factor of interactions (quarantine) of infected-aware individuals. Symptomatic individuals are already assumed to have a self or imposed partial precautionary element of quarantine even if unaware of infection, this is described by the parameter (**rfi**_**s**_). Also, in asymptomatic and mild disease individuals tested, the sensitivity of the tests is around 80%. In other words, of every 100 infected individuals tested, only 80 will yield a positive test and 20 will be false negative tests.

The awareness of infection after a positive test is assumed to lead to a full quarantine and removes those individuals from regular interaction with others. The fractions of individuals (without and with symptoms) aware of infection is therefore equal to the product of the fraction of total individuals randomly tested from the entire population (pt_ps_ for PS) and from the entire pool of individuals showing symptoms (pt_s_ for S) times the corresponding test sensitivity (t_sns_ps_ and t_sns_s_ respectively). Different types of tests may be available or adequate for each of the two groups.

The fractions of infectious PS and S individuals that remain in interaction with others (*f*_ips_ and *f*_is_) are therefore calculated as per Eqs 5.e-f. Hospitalised, critical and deceased are considered excluded from the pool of interacting individuals.

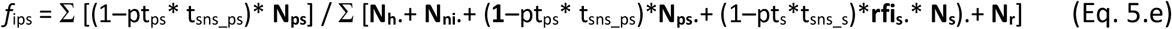

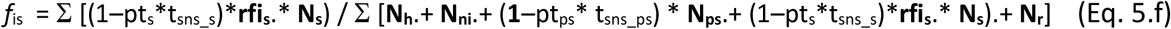

where pt_s_ is the proportion of symptomatic individuals tested (randomly) and pt_ps_ is the proportion of randomly tested non-symptomatic individuals of all types. For symptomatic S individuals both RT-qPCR and serological tests are assumed possible while, for non-symptomatic, only RT-qPCR tests. The parameters t_sns_s_ and t_sns_ps_ refer to the sensitivity of the tests for both groups respectively. Table 5 shows the definitions and units of all the parameters used in the modelling of interventions.

#### Critical care capacity

The impact of available critical care capacity is modelled by a specific function to allocate critically ill individuals as per the available ICU. The function allocates critically ill individuals in two possible groups, namely those admitted to ICU (**N**_**sc_ic**_) and those not admitted to ICU due to lack of capacity or for medical or humanitarian reasons (**N**_**sc_ncc**_). At each simulation time point the allocation function is computed for the total **N**_**sc**_ per age group.

The function allocates ICU resources with priority to age groups with higher ICU survival rate (f_r_sc_) until the maximum number of intensive care units is reached leaving any remaining individuals without care, in this way **N**_**sc_ic**_ and **N**_**sc_ncc**_ are computed.

As the COVID-19 outbreak has progressed, data indicate that not all patients in critical condition have been admitted into intensive care units (ICU). Data show that many individuals with very poor prognosis, particularly those of oldest age may have never been referred to ICU due to capacity limitations or other medical humanitarian reasons. This is based on data from Spain (Ministerio Sanidad España: Act. 107 COVID-19) showing that for individuals over 70, only a fraction of the reported fatalities previously hospitalised was ever admitted to ICU and this was not due to ICU lack of capacity. In order to maintain consistency with the reported data (Ministerio Sanidad España: Act. 107 COVID-19) the parameters of f_d_sc_ and f_sc_sh_ have been estimated such that the product of f_d_sc_ * f_sc_sh_ (fatality ratios over hospitalised individuals) is consistent with reported numbers for all ages irrespective of reported ICU admissions.

### Stage transition equations

The dynamic variation on the number of individuals in each stage over time and per age group is governed by the population balance equations described in Eqs 5.a-h. (all in vectors by age group).

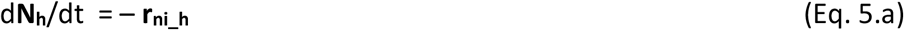

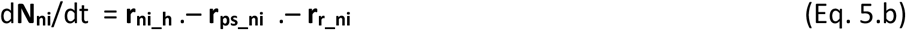

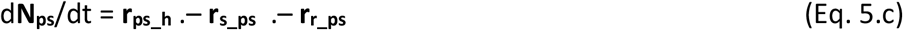

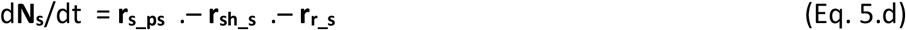

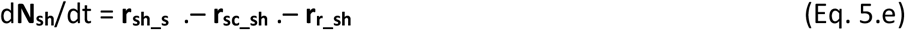

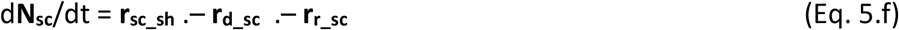

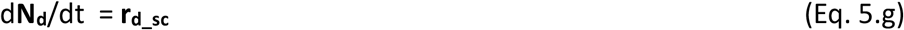

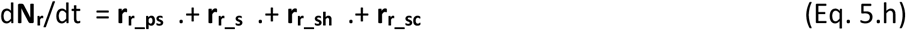

The state transitions as governed by these rates are represented in a matrix form in Figure S2.

### Calculation of the dynamic reproduction number (R_t_)

The reproduction number refers to the potential infection of susceptible individuals from infected individuals (Delamater *et al*. 2019). Since the model produces, for each parameters set used, a unique deterministic set of values for its outputs over at any given time, an instantaneous deterministic estimation of the reproduction number (R_t_) is also obtained. Several parameters and variables influence the R_t_ including the duration of infectious stages; the likelihoods of infection per social contact as well as the percentages of individuals transitioning to more severe stages.

The dynamic reproduction number (R_t_) during the outbreak is computed over time according to Eq. 6 from the current values of the model state variables. Under this approach, infectious individuals can only infect others while they are in pre-symptomatic (PS) and symptomatic (S) stages. Although it has been speculated that post-symptomatic recovered individuals may be infectious for some period of time, this has not been considered in the model at this time due to lack of data. Hospitalised and critical individuals are assumed to be well isolated and also not able to infect others. The provided dynamic output of the reproduction number R_t_ can be used to guide and interpret the impact of interventions in terms of R_t_.

Modelled infected individuals can take only three possible infectious paths, namely: (i) PS ⟶ R; (ii) PS ⟶ S ⟶ R and (iii) PS ⟶ S ⟶ SH. These paths are made of combinations of four possible infectious stage intervals in which infected individuals spend time and infect at their corresponding rate (see Table 6).

**Table 6.** Possible infectious stages intervals for R_t_ computation.

| Infectious interval | Fraction of infected passing the interval<br>($\text{ind}_{\text{interv}}/\text{ind}_{\text{inf}}$) | Interval duration<br>(d) | Total infections per stage interval per individual infected<br>($\text{inf}_{\text{interv}}/\text{ind}_{\text{inf}}$) |
| --- | --- | --- | --- |
| $PS \rightarrow R$ | $f_{r\_ps}$ | $t_{r\_ps}$ | $(r_{i\_psT} / N_{psT}) * t_{r\_ps} * f_{r\_ps}$ |
| $PS \rightarrow S$ | $(1 - f_{r\_ps})$ | $t_{s\_ps}$ | $(r_{i\_psT} / N_{psT}) * t_{s\_ps} * (1 - f_{r\_ps})$ |
| $S \rightarrow R$ | $(1 - f_{r\_ps}) * f_{r\_s}$ | $t_{r\_s}$ | $(r_{i\_sT} / N_{sT}) * t_{r\_s} * (1 - f_{r\_ps}) * f_{r\_s}$ |
| $S \rightarrow SH$ | $(1 - f_{r\_ps}) * (1 - f_{r\_s})$ | $t_{sh\_s}$ | $(r_{i\_sT} / N_{sT}) * t_{sh\_s} * (1 - f_{r\_ps}) * (1 - f_{r\_s})$ |

The dynamic computation of R_t_ consists of adding the total infection contributions of every stage interval as shown in Eq. 7.

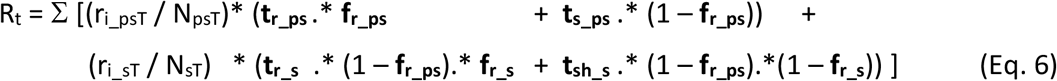

in which, the age group weighted average rates of infection by PS and S are given as per Eqs. 6a-b.

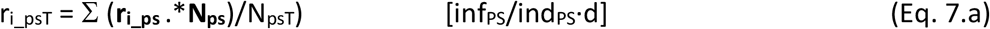

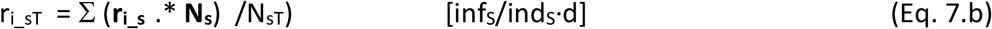

### Model limitations

The model shares many of the fundamental characteristics of compartment SIR-type models and is based on dynamic balances of individuals in compartments classified by their stage of infection and age groups only, no other differentiation within those groups is captured by the model. This characteristic allows for the model application only to single, densely populated clusters. The model has a low complexity and requires a small number of parameters that are also mechanistic and meaningful. Most of these parameters can be directly estimated from epidemiological and clinical data and are not recommended for calibration against data. Analogous to all SIR-type models this model carries limitations since all variables and parameters refer to representative averages for each compartment of stage and age group. This may limit the model representation of the non-linear interactions that occur in reality. Phenomena like so-called super spread events or any location specific phenomena are not reproduced by these types of models. Any quantitative interpretation of results for prediction purposes should therefore be always accompanied by a critical discussion against these limitations.

## Impact of interventions on a COVID-19 outbreak case study

A case study based on a scenario of propagation of the COVID-19 pandemic using data available as of May 2020 is presented below. The results obtained are intended to be interpreted qualitatively and to be contextualized to the specific setting characteristics. They serve also as a demonstration of the model potential if applied with higher confidence parameter values. Several selected scenarios were simulated aimed at illustrating the impact of different interventions.

Default reference epidemiological and clinical parameter values were extracted from different information sources on the COVID-19 outbreak as available in May 2020. Details of values and sources are provided in the Supplementary Information Tables S1-S2 respectively, with indications of the level of confidence. A population with an age distribution as that of the region of Madrid (Spain) in 2019 was used (INE Spain, 2020) during the simulations.

Default reference values for interventions-related parameters were selected arbitrarily for a situation assimilated to that previous to the outbreak and without any specific intervention (see values and rationale in the Supplementary Information Table S3). The value for available intensive care beds per million people (capICpM) of 261 has been used by default in all case studies. The dynamic simulation results of the default outbreak scenario under no intervention is shown in the Supplementary Information Figure S4.

All scenarios are simulated for 365 days and evaluated in terms of (i) final total number of fatalities at outbreak termination and (ii) final number of fatalities per age group. In addition, the scenarios are presented also in terms of dynamic profiles over time for (iii) number of active cases; (iv) reproduction number; (iv) number of critical cases; (v) number of fatalities.

### Intervention #1. Social isolation

In this scenario, the impact of the level of imposed social isolation was evaluated by age brackets. Four cases are shown, namely universal social isolation, that for elderly only, youngsters only, and elderly and youngsters only.

The parameter that describes this intervention is the reduction in the number of interpersonal contacts that healthy susceptible individuals have with others per day (**ni**_**h**_). Figure 4 shows the comparison between the impacts of the four types of isolations. The Supplementary Information Appendix VI contains the complete results of the four isolation intervention types.

**Figure 4.**
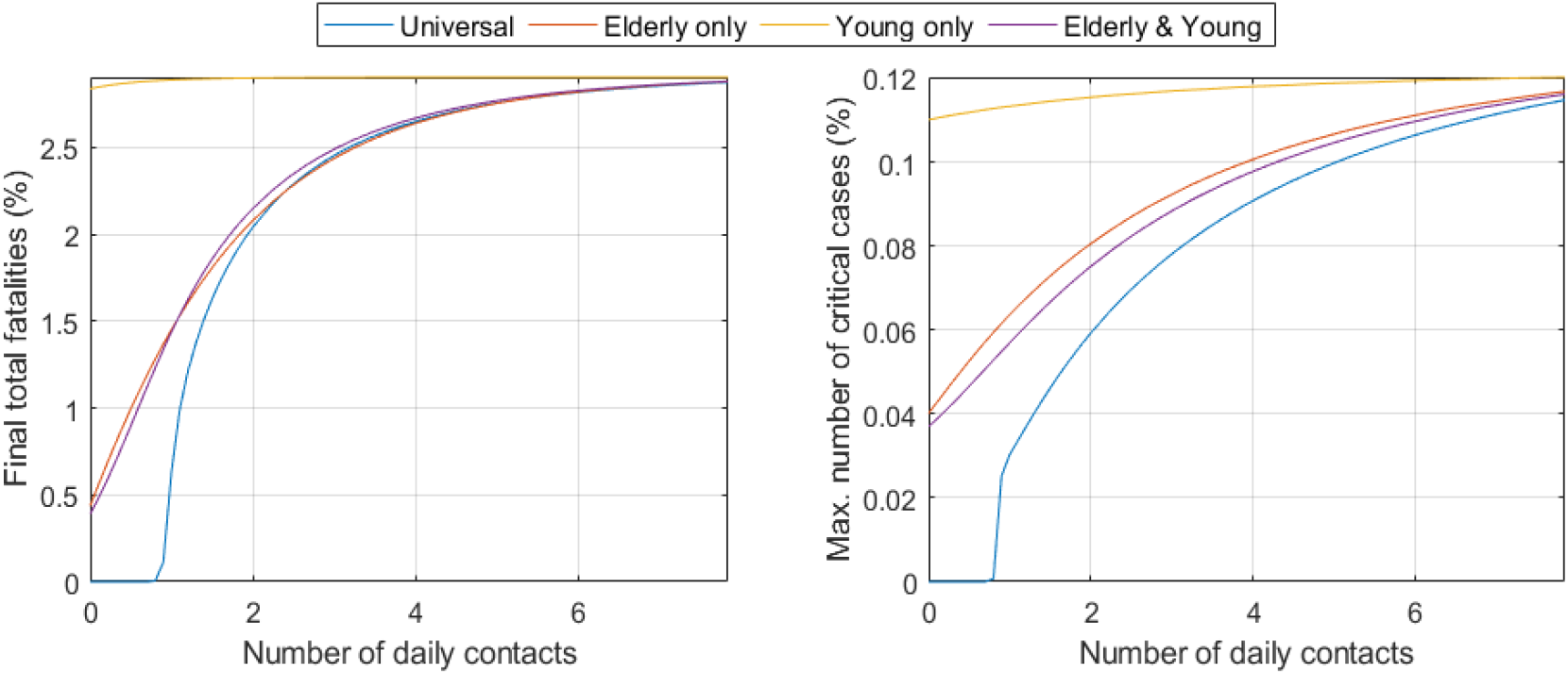
Impact of the level of social isolation under the four different strategies universal and per age groups on the total number of fatalities at the end of the outbreak (left) and on the maximum critically ill individuals ever reached during the outbreak (right). Numbers are as percentage of the total population.

It appears that the intervention starts to be effective in terms of significantly reducing total deaths after the number of daily contacts is placed below a threshold number, above which there is minor impact. Interestingly, the ni_h_ does not appear to significantly modify total final mortality beyond one single contact per day. The number of fatalities appears clearly and directly related to social isolation as well as the speed at which the fatalities saturation will occur. Full details can be seen in Figures S6. The model is capable of capturing this partly due to its description of the saturation of the healthcare capacity and withdrawal of critical care over capacity.

The middle and bottom graphs in Figure S6.a show the impact of ni_h_ on the time course of several variables. Figure S6.a (middle left) supports the “flatten the curve” concept, now globally popular. If interactions are not modified, the number of cases grows rapidly explosively. Figure S6.a(middle right) shows the estimate of R_t_ over time. This illustrates how the level of social isolation can define the infectability and the number of cases each infected individual will infect (R_t_) showing how factors such as interventions can impact R_t_. The number of critical cases increases throughout time as the social interaction increases Figure S6.a(bottom left).

The detailed results of imposed social isolation selective to those over 60 years old is shown in Figure S6.b. As shown in Figure 4 the selective social isolation of the elderly has a potentially very significant impact on final total fatalities at an almost comparable level than for universal isolation. This is a result with potentially significant consequences as it indicates that a sustained isolation selective only to the elderly and not to the other age groups could alleviate the economic damage at a cost of a small number of increased total fatalities. The decrease in social interactions in schools and colleges by isolation of the young may however have an impact on the overall multiplier of infections from youngsters to adults.

The impact of selective imposed social isolation of only those under 20 years old was also evaluated. Figure S6.c shows the results for this scenario, for the output variables indicated, in absence of other interventions. The young population have been observed to be quite resistant to the disease. The isolation of the young produces no effect in the overall final fatality rate but produces a moderate impact on the mortality of the elderly at low values of ni_h_. As it can be seen in Figure 4, social isolation of the young has little impact producing almost identical curves for any levels of social isolation. It is thought however that the decrease in social interactions in schools and colleges by isolation of the young may have a large impact on the overall multiplier of infections from youngsters to adults. This emergent aspect of the disease spread behaviour and containment efforts is captured in our results, even though the present model does not incorporate geographical features and does not explicitly describe location-specific population interactions (such as those synthetic location-specific contact patterns in *Prem et al*., 2020).

The fourth case of selective imposed social isolation for both those under 20 and those over 60 years old is also evaluated. The number of daily social contacts with other people that healthy susceptible individuals of the age groups under 20 and over 60 years old have (**ni**_**h**_) is modified. The full results for this scenario are shown in Figure S6.d.

Many of the early interventions during the COVID-19 outbreak started by protecting the elderly and isolating the young (no schools, no colleges or universities for students), decreasing the number of interactions of the two subpopulations substantially. The isolation of these population groups together results in similar effects to that of the isolation of the elderly alone with no significant added value in isolating the young respect to that the elderly alone as shown in Figure 4.

### Intervention #2. Use of PPE and safety distance

The impact of increased use of PPE and social distancing awareness is evaluated in this scenario. The parameter that describes this intervention is a factor increasing the default values (see Table S3) of the level of personal protection and awareness (**lpa**) parameters of the healthy and infected population groups (**lap**_**h**_, **lap**_**ps**_ and **lpa**_**s**_). Increases in these parameters decrease the likelihood of infection per interaction (see Eqs. 4) and subsequently the rates of infection (Eqs. 5). Figure 5 presents the main impacts of on this intervention on the total final fatalities and peaks of critical cases. More complete results, including the predictions of the dynamic reproduction number R_t,_ are presented in Figure S7.

**Figure 5.**
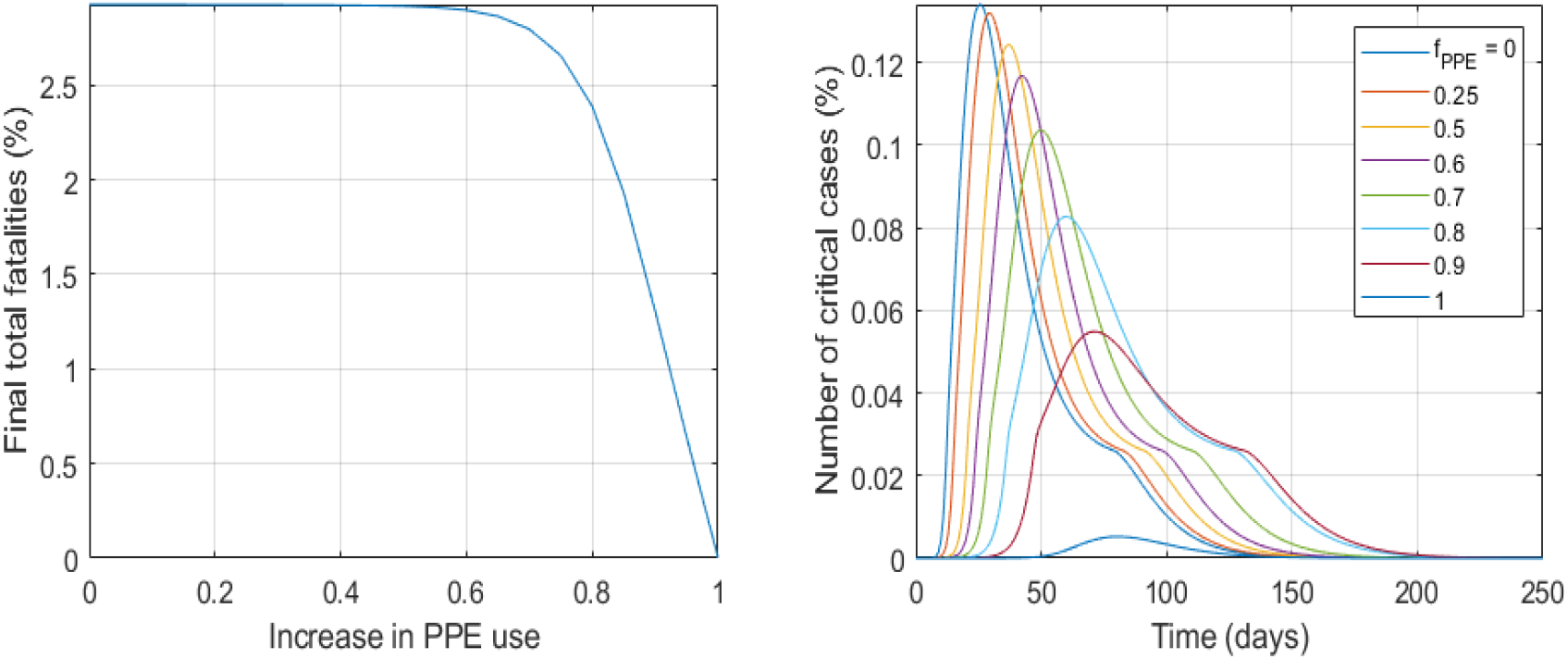
Impact of the level of PPE use and social distancing on the total number of fatalities at the end of the outbreak (left) and on the peaks of critically ill individuals (right). Numbers are as percentage of the total population.

As it is shown in Figure 5 the extensive use of PPE appears as having a potentially major impact on total outbreak fatalities at the highest levels of protection. There is an inverse relationship between the level of protection and the overall fatality of the disease. The peak of number of cases is reached earlier and is higher if low levels of personal protection are used, the infectability and R_t_ follow the same pattern. The height of the peak number of critical cases is much lower is extensive use of PPE is implemented as seen in Figure 5 (right), if the peak does not exceed the critical care capacity the total number of fatalities reaches much smaller values.

### Intervention #3. Awareness of infection by testing

The impact of widespread extensive testing is evaluated in this scenario. The effect of the percentage of detected infections on the final total fatalities and outbreak dynamics is evaluated. The compared impact of testing only symptomatic individuals, testing randomly only the non-symptomatic population (therefore the same fraction of pre-symptomatic) and testing everyone both with and without symptoms. The fractions of infections detected in each group (S and PS) is the product of the fraction of the group tested pt_s_ or pt_ps_ respectively times the sensitivity of each group’s test t_sns_s_ and t_sns_ps_ (different test sensitivities for each type of test apply).

The model describes the impact of infection detection as a reduction in the fraction of infectious (PS and S) individuals available for interactions among the total ones. This due to their knowledge of infection and subsequent (self)quarantine. See Eq. 5.e-f. These reduced fractions of infectious individuals among total reduces the rates of infection by both groups (Eq. 4.a-b).

If only symptomatic individuals are tested, the impact is almost negligible and only when both S and PS groups are extensively tested to levels allowing for a detection of near 90% of total infections a meaningful impact is predicted. The current test sensitivities (around 80%) imply that even at 100% of population tested those high levels of detentions required will be unachievable and a large number of infections will remain undetected yielding the intervention ineffective.

The results clearly show that extensive testing appears as a non-effective intervention due to the unreachable very high percentage of infection detections required for it to have an impact (see Figure 6). Figures S8.a-c. show the complete simulation results of the impact of this intervention.

**Figure 6.**
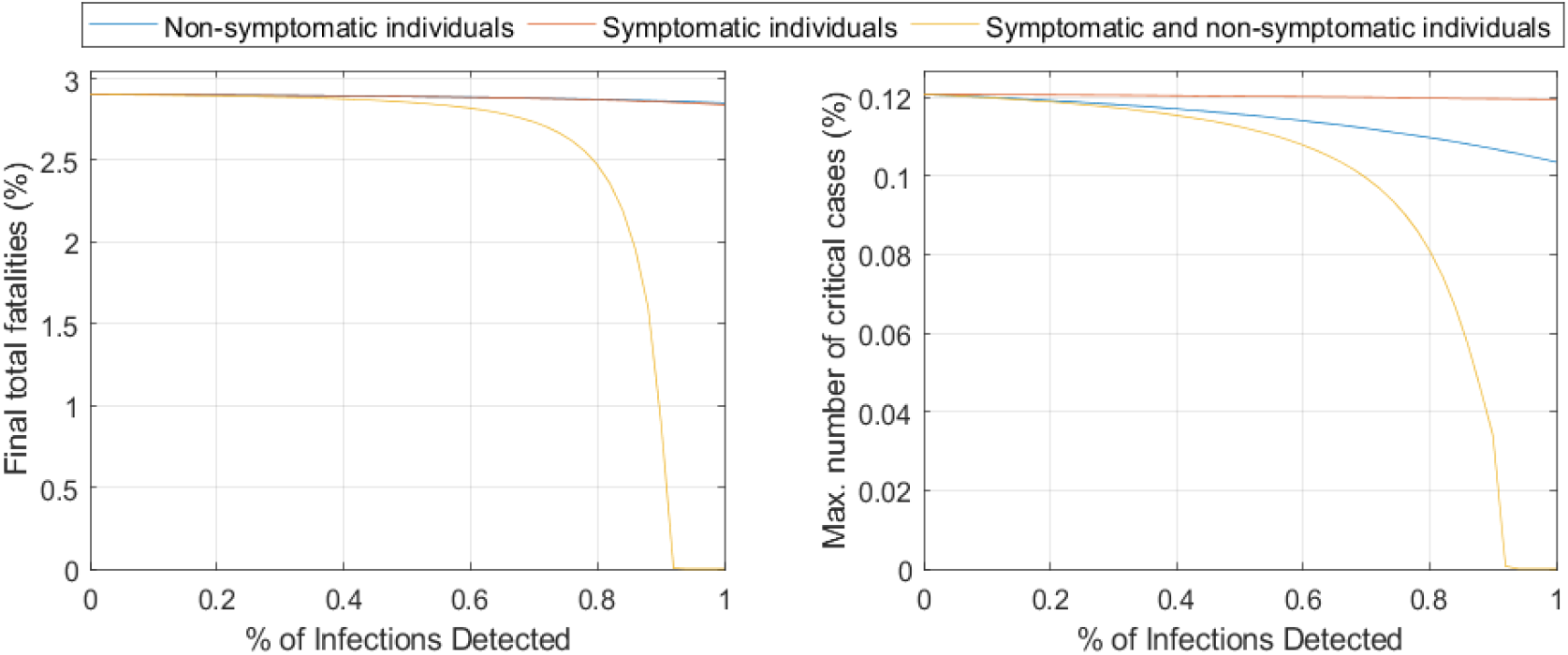
Impact of the infection detection level (product of the proportion of tested individuals times the test the sensitivity, 80%) for symptomatic alone, pre-symptomatic alone and for both. The impact on the total number of fatalities at the end of the outbreak (left) and on the maximum values of critically ill individuals ever reached during the outbreak (right) are shown. Numbers are as percentage of the total population.

### Intervention #4. Critical care capacity

The impact of the availability of intensive care beds is evaluated in this scenario. The parameter that describes this intervention is the number of available intensive care beds per million population. Figure 7 illustrates the model predictions for this scenario, in terms of total final fatalities and numbers of critical cases over time. Note that once the ICU beds capacity is exceeded the critically ill patients become fatalities in one day.

**Figure 7.**
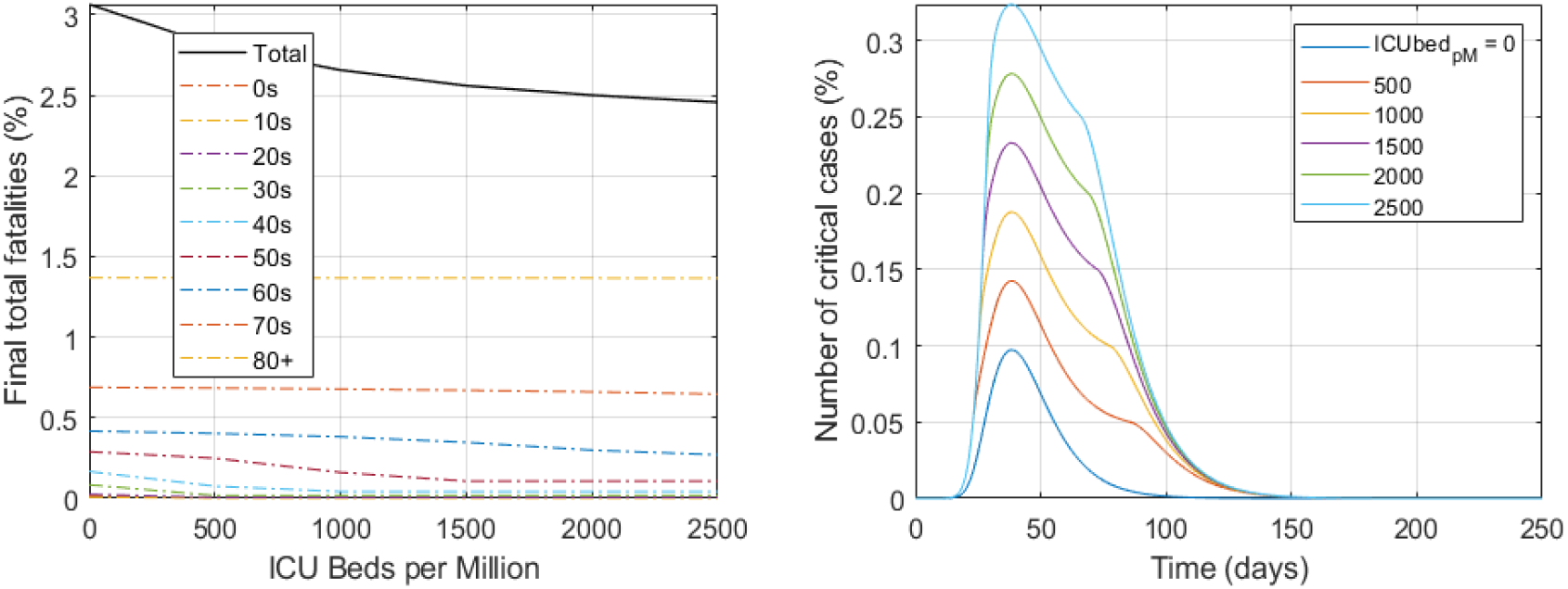
Impact of the availability of intensive care beds on the final number of fatalities total and also per age group (left) as well as for the different time course profiles of the number of critical cases (right). Numbers are in percentage of the total population of all ages.

From Figure 7 (and Figure S9 for more complete details), the enormous impact that the increase in critical care resources can have in decreasing total fatalities becomes evident. The higher the availability of critical beds, the lower number of fatalities, the trend applies until there is no shortage of IC beds and all remaining fatalities are only the unavoidable ones. This intervention avoids those deaths that are preventable by the availability of critical care support for those that need it.

## Conclusions on the impact of interventions

The impact of specific interventions on the outbreak time course, number of cases and outcome of fatalities were evaluated. Data available from the COVID-19 outbreak as of May 2020 was used. Our results on the impact and mechanism of several interventions indicate that:

1. Universal social isolation measures may be effective in reducing total fatalities only if they are strict and the average number of daily social interactions is reduced to very low numbers. Selective isolation of only the age groups most vulnerable to the disease (i.e. older than 60) appears almost as effective in reducing total fatalities but at a much lower economic damage.
2. The use of protective equipment (PPE) appears capable of very significantly reducing total fatalities if implemented extensively and to a high degree.
3. Extensive random testing of the population leading to infection recognition and subsequent immediate (self) isolation of the infected individuals, appears to be an ineffective intervention due to the required (unreachable with existing test sensitivities) high percentage of infection detections and the incapability to be sustained over time.
4. An increase in the number of critical care beds to directly save significant numbers of lives with a direct reduction in total final fatalities per each extra available critical care bed unit.

It is important to note that any quantitative interpretation of the results must be accompanied with a critical discussion in terms of the model limitations and its frame of application. The sensitivity analysis provided should be used to help such analysis.

## Conclusions on model application and roadmap for model expansion

The confidence in the model is based on the confidence of its epidemiological and clinical parameters. As time passes the quality of data towards this end is increasing rapidly. Any calibration of epidemiological or clinical parameters against dynamic data curves is not recommended as it will likely lead to model overfitting. Instead, valid epidemiological and clinical sources should be used for estimation, and not calibration, of these parameters.

Only the intervention parameters are recommended for calibration using dynamic data curves and should be conducted against data from representative populated cities or well mixed communities. Data from cities in which population is typically interconnected socially in public areas and public transport is widely used and are particularly well suited to the calibration of these parameters. The use of dynamic data for entire countries with regions in which the outbreak may be in different stages is not recommended. The recommendation then is to fit the model to each independent, similar population. This type of model does not represent well the outbreak dynamics unless the population is well mixed and in the same outbreak stage.

More detailed descriptions and sub models of some of the intervention relevant parameters such as the levels of social interaction and personal protection measures are avenues for model further development.

The model modularity and its fast computation allows for its easy scale up into multiple population nucleus that could be simulated in parallel with degrees of interconnectivity among them. Separate independent copies of the model can be run in parallel one for e.g. each city in a region or country and migration terms can be added between cities. Interventions can then be defined to include e.g. travel restrictions between those cities at different levels.

The mechanistic nature of the model makes it also very suitable for the evaluation of advanced optimisation and optimum control strategies. Its capacity of describing complex interactions makes it also of potential use to develop advanced artificial intelligence (AI) algorithms to aide and provide advice to authorities during decision making. AI algorithms could be trained by evaluation of very large numbers of scenarios combining static and dynamic interventions of different types against total fatalities and economic damage.

## Additional Information and source code

The for Matlab® source code and Excel file containing all parameter values used as well as a non-age segregated version of the model are available at https://github.com/EnvBioProM/COVID_Model

## Data Availability

The for Matlab source code and Excel file containing all parameter values used as well as a non-age segregated version of the model are available at https://github.com/EnvBioProM/COVID_Model

## Acknowledgements

All authors wish to thank Khalifa University (Grant 8474000317 CRPA-2020-SEHA) and the Government of Abu Dhabi for the funding and support.

## Notes

### Competing Interest Statement

The authors have declared no competing interest.

